# Cross-sectional associations between 24-hour time-use composition, grey matter volume and cognitive function in healthy older adults

**DOI:** 10.1101/2023.05.15.23289982

**Authors:** Maddison L Mellow, Dorothea Dumuid, Timothy Olds, Ty Stanford, Jillian Dorrian, Alexandra T Wade, Jurgen Fripp, Ying Xia, Mitchell R Goldsworthy, Frini Karayanidis, Michael J Breakspear, Ashleigh E Smith

**Affiliations:** Alliance for Research in Exercise, Nutrition and Activity, Allied Health and Human Performance, University of South Australia, Australia; Behaviour-Brain-Body Research Centre, Justice and Society, University of South Australia, Australia; The Australian e-Health Research Centre, CSIRO Health and Biosecurity, Brisbane, Queensland, Australia; School of Biomedicine, University of Adelaide, Australia; Hopwood Centre for Neurobiology, Lifelong Health Theme, South Australian Health and Medical Research Institute (SAHMRI), Australia; Functional Neuroimaging Laboratory, School of Psychological Sciences, College of Engineering, Science and the Environment, University of Newcastle, Australia; Discipline of Psychiatry, College of Health, Medicine and Wellbeing, University of Newcastle, Callaghan, NSW, Australia

**Keywords:** time use, physical activity, sleep, sedentary behaviour, brain volume, cognitive function, aging

## Abstract

**Background:** Increasing physical activity (PA) is an effective strategy to slow reductions in cortical volume and maintain cognitive function in older adulthood. However, PA does not exist in isolation, but coexists with sleep and sedentary behaviour to make up the 24-hour day. We investigated how the balance of all three behaviours (24-hour time-use composition) is associated with grey matter volume in healthy older adults, and whether grey matter volume influences the relationship between 24-hour time-use composition and cognitive function.

**Methods:** This cross-sectional study included 378 older adults (65.6 ± 3.0 years old, 123 male) from the ACTIVate study across two Australian sites (Adelaide and Newcastle). Time-use composition was captured using 7-day accelerometry, and T1-weighted magnetic resonance imaging was used to measure grey matter volume both globally and across regions of interest (ROI: frontal lobe, temporal lobe, hippocampi, and lateral ventricles). Pairwise correlations were used to explore univariate associations between time-use variables, grey matter volumes and cognitive outcomes. Compositional data analysis linear regression models were used to quantify associations between ROI volumes and time-use composition, and explore potential associations between the interaction between ROI volumes and time-use composition with cognitive outcomes.

**Results:** After adjusting for covariates (age, sex, education), there were no significant associations between time-use composition and any volumetric outcomes. There were significant interactions between time-use composition and frontal lobe volume for long-term memory (p=0.018) and executive function (p=0.018), and between time-use composition and total grey matter volume for executive function (p=0.028). Spending more time in moderate-vigorous PA was associated with better long-term memory scores, but only for those with smaller frontal lobe volume (below the sample mean). Conversely, spending more time in sleep and less time in sedentary behaviour was associated with better executive function in those with smaller total grey matter volume.

**Conclusions:** Although 24-hour time use was not associated with total or regional grey matter independently, total grey matter and frontal lobe grey matter volume mediated the relationship between time-use composition and several cognitive outcomes. Future studies should investigate these relationships longitudinally to assess whether changes in time-use composition correspond to changes in grey matter volume and cognition.

## 1. Background

Healthy ageing is associated with changes in cortical volume and cognitive function (1, 2). For example, older adults without diagnosed cognitive impairment may still present with cognitive challenges in vocational and interpersonal domains. Effective strategies which slow reductions in brain volume may have additional benefits for maintaining healthy cognitive functioning in older age and preventing or delaying future cognitive decline and dementia. One strategy is through achieving and maintaining sufficient physical activity. Physical activity has been identified as a modifiable risk factor for dementia in older adulthood (3) and is positively associated with both brain volume and cognitive function in healthy older adults. Cross-sectional and longitudinal magnetic resonance imaging (MRI) studies suggest that older adults who engage in higher levels of PA have greater global brain volume (4–8) and greater volume in medial temporal (9, 10) and frontal regions (11). Similarly there is evidence that habitual physical activity is positively associated with cognitive function in healthy older adults across several cognitive domains (12). The mechanisms underlying this relationship are not well understood, however it is likely that slowing reductions in cortical volume contributes to the positive association between physical activity and cognitive function.

Although physical activity is positively related to both brain volume and cognitive function in older adults, an important but often overlooked consideration is that physical activity does not take place in isolation. Physical activity must fit within the 24-hour day. In fact, the 24-hour day can be broadly divided into three time-use behaviours: physical activity, sedentary behaviour, and sleep, which are each related to brain health. Sedentary behaviours, defined as time spent in a sitting or reclined position and expending <1.5 metabolic equivalents (METs), may impact brain health through mechanisms that differ from those involved in physical inactivity (13). To date, few studies have investigated the relationship between sedentary behaviour and grey matter volume in older adults. A recent study found no differences in total grey matter volume between high and low sedentary behaviour groups (<8 hours or >8 hours per day), but reported a trend of lower hippocampal volumes in the high sedentary behaviour group (14). Two additional studies reported a relationship between high levels of sedentary behaviour and cortical thinning in hippocampal sub-regions (i.e., parahippocampal cortex, entorhinal cortex, and subiculum) (14, 15). Similarly, the associations between sedentary behaviour and cognitive function in older adults are not clear (12). Several mechanisms, including modulation of cerebral blood flow, neurotrophic factors, and brain structure (16, 17), have been postulated to mediate links between sedentary behaviour and cognitive function. However, it is hypothesized that grey matter volume may be less sensitive to the cardiovascular health effects imposed by excessive sedentary behaviour compared to other aspects of brain structure such as the volume and composition of white matter (18).

There is also mixed evidence on associations between sleep, brain structure and cognitive function in older adults. Some studies have indicated that short or long sleep durations (i.e., <6HR or >9hr per night) are associated with loss of cortical volume in frontal and temporal regions (19) and ventricular enlargement (20) in older adults. Similarly, short or long sleep duration has been negatively associated with cognitive function in older adults (21, 22). Conversely, a recent 28-year longitudinal study found no significant differences in grey matter volume or cognitive changes between older adults who slept for 5, 6, 7 or 8 hours per night. However, the null findings in that study may have been driven by the small number of participants in ‘extreme’ groups (i.e., most participants slept 6-7 hours per night which more closely aligns with sleep guidelines) (23). It is not well understood whether poor sleep causes reduced cortical volume, whether reduced cortical volume causes poor sleep, or whether the relationship is bi-directional (24). In summary, the mechanisms underlying the relationship between sleep and cognitive function in older age remain unclear, and it is hard to disentangle direct effects from a wide range of possible confounders.

To date, physical activity, sleep, and sedentary behaviour have been studied independently in relation to grey matter volume, or while considering only two of the three behaviours together. However, physical activity, sleep and sedentary behaviour interact to make up the 24-hour day, such that increasing time in one behaviour will lead to a reduction in one or both remaining behaviours (25). It is therefore more meaningful to consider the effects of time-use composition (or the proportion of time spent in each time-use behaviour within the 24-hour period) on brain structure. A recent study by Maasakkers et al. (26) found that sedentary participants had smaller hippocampal volumes, but this relationship was attenuated when controlling for levels of moderate-vigorous physical activity (MVPA). Thus, the interactions between time use behaviours may be important for grey matter volume beyond the independent effects of each time-use behaviour alone.

Understanding the interactive effects of physical activity, sedentary behaviour and sleep on grey matter volume may help inform 24-hour movement guidelines for healthy ageing and dementia prevention in older adults. Further, understanding whether the association between 24-hour time-use composition and cognitive function is mediated by grey matter volume will provide further insight into the mechanisms by which optimal time use benefits cognitive functioning in older adulthood. To our knowledge, no studies have investigated how 24-hour time-use composition is associated with grey matter volume in healthy older adults. Our previous study found weak associations between independent time-use behaviours (but not 24 hour time-use composition) and cognitive function in a similar sample of healthy older adults (27) but did not explore neurophysiological mechanisms underlying these relationships. To address these important gaps in knowledge, cross-sectional data from the baseline phase of the ACTIVate study were used to investigate (a) whether 24-hour time-use composition is associated with grey matter volume, and (b) whether grey matter volume mediates the relationship between 24-hour time-use composition and cognitive function.

## 2. Methods

### 2.1 Ethics

The ACTIVate study was registered with the Australia New Zealand Clinical Trials Registry (ACTRN12619001659190) on November 27, 2019. Ethics approval was obtained from the University of South Australia and University of Newcastle Human Research Ethics Committee (202639). All procedures were conducted in accordance with the Declaration of Helsinki.

### 2.2 Participant recruitment and screening

Eligibility criteria for the ACTIVate study have been reported in detail elsewhere (28). Briefly, participants were eligible if they were aged 60-70 years, fluent in English, had no clinical diagnoses of dementia or any other neurological or psychiatric disorders, did not have an intellectual or major physical disability, and presented no contraindications to transcranial magnetic stimulation or MRI screening tools (29).

Participants were recruited for the ACTIVate study using a rolling convenience sampling strategy (28). Those who met initial eligibility criteria were further screened against cognitive impairment using the Montreal Cognitive Assessment (blind) via phone interview. Participants who scored <13 (out of a potential 22) were excluded from the study.

Power calculations were used to determine the required sample size for the larger ACTIVate study (based on cognitive outcomes), which have been detailed extensively elsewhere (28). Briefly, aiming for 80% power, allowing for attrition and response rate at recruitment and accounting for the longitudinal design of the study, the final sample size of 448 participants was determined.

### 2.3 Study measures

#### 2.3.1 Device-measured time-use patterns

Data were collected between August 2020 and February 2022. Measures of daily time-use patterns (time spent in physical activity, sedentary behaviour, and sleep) were derived from triaxial accelerometers. Participants wore an Axivity AX3 monitor on their non-dominant wrist for 7 consecutive days and were asked to complete a sleep log for each day of wear (time they got out of bed, time they went to bed, time spent napping during the waking day, and reasons for removal of monitor during the day). Accelerations were sampled at 100Hz. Raw acceleration data were downloaded using the Open Movement GUI software (OmGUI; Newcastle, UK), converted to .CSV files and processed using a custom MATLAB graphic user interface developed by researchers at the University of South Australia (COBRA; MATLAB R2018B).

Time spent in sleep was classified manually by marking the wake and sleep times for each 24-hour recording period while cross-checking participants’ sleep logs against the accelerometry trace. Brief nighttime awakenings were not captured unless waking periods were explicitly reported by participants. Waking day behaviours were classified as time spent in MVPA (>93 g-min), light intensity PA (LPA; >48 g-min) or sedentary behaviour (<48 g-min) using previously published cut points adjusted for sampling frequency (30). ‘Valid wear days’ were classified if accelerometers were worn for at least 10 waking hours. To be included in final analyses, participants were required to have at least three valid weekdays and one valid weekend day. Total time spent in each time-use behaviour was averaged across the recording period, providing values reflecting the average time spent in MVPA, LPA, sedentary behaviour, and sleep per day (in minutes).

#### 2.3.2 Brain imaging and MRI processing

MRI acquisition was performed on a Siemens Skyra 3T scanner in Adelaide, and a Siemens Prisma 3T scanner in Newcastle, both using a 64-channel head and neck coil. T1-weighted magnetization prepared rapid gradient echo (MPRAGE) images were acquired for volumetric quantification of brain structures, and T2-weighted fluid-attenuated inversion recovery (FLAIR) images were used to assess white matter hyperintensity burden. The acquisition parameters for MRI sequences are described elsewhere (28).

3D T1 MPRAGE images were first segmented into grey matter, white matter and cerebrospinal fluid tissues using an in-house implementation of the expectation maximization algorithm (31). The brain parcellation was performed using the NeuroMorphometrics parcellation atlas and Learning Embeddings for Atlas Propagation method (32), allowing measurement of cortical and subcortical grey matter volumes. White matter hyperintensities were quantified from T2 FLAIR images using the HyperIntensity Segmentation Tool (33). For the purpose of this study, white matter hyperintensity volume data were only used to characterize the sample and were not included in main analyses.

Volumetric measures of grey matter regions of interest (ROIs) were derived from the brain segmentation and the NeuroMorphometrics parcellation, which included total grey matter, lateral ventricle, bilateral frontal lobe, bilateral temporal lobe and bilateral hippocampus volumes.

#### 2.3.3 Cognitive function measures

Cognitive function was measured using a series of tests from the Cambridge Automated Neuropsychological Test Automated Battery (CANTAB). Using the Cattell-Horn-Carroll-Miyake taxonomy (34) as a guiding framework, tests were z-scored and then combined to create three cognitive composites: long-term memory (Verbal Recognition Memory test); executive function (Multitasking and One Touch Stockings of Cambridge tests); and processing speed (Reaction Time test). The average z-score of cognitive tests within each composite (i.e., a single z-score value for each cognitive domain) was used in final statistical models. Higher z-scores indicated better cognitive performance. The methods used to create these composites have been described in detail elsewhere (27).

#### 2.3.4 Covariates

Age (years), sex (male, female) and education (total years) were entered as covariates in linear regression models, based on previous evidence of their associations with grey matter volume (35–37) and cognitive function (3, 38). Age and sex data were derived from a demographics questionnaire, whilst total years of education (including primary, secondary and tertiary education) was derived from the Australian National University Alzheimer’s Disease Risk Index collected for the larger study (39).

### 2.4 Statistical analysis

All inferential statistics were conducted in R version 4.2.2 and the code used for data analysis is available at https://github.com/MaddisonMellow/time-use-brainvol-paper. To account for total brain size and MRI scanner/protocol differences, all ROI volumes were adjusted for total intracranial volume, scanner site (Adelaide or Newcastle), and use of the distortion correction option during scanning (on or off) using linear regression models. Next, outcome variables (ROI volumes) were inspected for normality and extreme skewness. At this stage of analysis, no transformations were performed as data were normally distributed, but data were further inspected and transformed later to improve model fit as needed.

#### 2.4.1 Pairwise correlations

To explore relationships between individual time-use variables (minutes/day in sleep, sedentary behaviour, LPA and MVPA), brain volume measures (total grey matter, lateral ventricle, frontal lobe, temporal lobe, and bilateral hippocampus volumes), cognitive outcomes (long-term memory, executive function, and processing speed) and continuous covariates (age and education), pairwise correlation coefficients were calculated. Pearson correlations were used for all pairwise correlations outside of the time-use composition (i.e., all correlations except for those *between* time-use behaviours). Because time-use behaviours are components of a composition, and therefore have a constant sum constraint (1440 minutes of the day), applying traditional correlation analysis between time-use variables may result in spurious correlations (40). Intuitively, because of the fixed sum constraint, an increase in one compositional part will result in lower values in remaining compositional parts which, when using a traditional Pearson’s correlation coefficient, would impose a negative correlation. To overcome this, symmetric balanced isometric log ratio coordinates are used (40). In essence, these symmetric balanced coordinates focus on the pairwise changes in two compositional parts (e.g., sleep and MVPA) compared to a representative value of the average of the remaining parts, using two sets of sensibly chosen isometric log ratio coordinates that are bisected (and length normalized) for each pairwise comparison of compositional parts. Thus, traditional correlation coefficients can be calculated on these transformed values, but the interpretation is not the strength of the linear relationship between the two variables/parts as with Pearson’s correlation coefficient. Rather, correlation coefficients of pairwise compositional parts expressed in symmetric balanced coordinates are interpreted as the “dominance” of one compositional part over another (similarly, values between -1 and 1): coefficients are positive when the two compositional parts increase simultaneously compared to a representative value of the average of the remaining parts, whereas coefficients are negative when the increase of one compositional part (e.g., MVPA) is associated with a decrease in the other compositional part (e.g., sleep) compared to a representative value of the average of the remaining parts. This method was applied to calculate correlations between time-use behaviours (i.e., within the 24-hour composition) using the corCoDa function in the *robCompositions* package (41).

#### 2.4.2 Compositional data analysis (CoDA)

All compositional analyses were conducted using the R *compositions* package version 1.4 (42). Daily time use compositions were created for each participant, representing the average proportion of time spent in MVPA, LPA, sedentary behaviour and sleep each day (summing 1440 minutes total). Time-use compositions were isometric log-ratio transformed to be included in statistical analyses as predictors (see Dumuid et al. (25) for overview of this method).

First, linear regression models were used to derive the associations between 24-hour time-use composition (predictor) and each ROI volume (outcome). Model 1 included covariates only (age, sex, education), whilst Model 2 included covariates and the predictor of interest (time-use composition). Model fit was examined using the *performance* package in R (43). To improve the model fit, lateral ventricle volumes were log transformed, whilst all other model fit diagnostics passed assumption checks and variables were therefore not transformed. To account for the possibility that associations between time-use composition and ROI volume outcomes may be non-linear (e.g., inverted U-shaped relationships between sleep and ROI volume), an additional model (Model 3) which was identical to Model 2 but expressed time-use composition using quadratic (squared) terms was fit. We used an F-test to explore whether quadratic terms (Model 3) improved model fit compared to Models 1 and 2. Expressing time-use composition using quadratic terms did not improve the model fit for any ROI volumes compared to the standard linear regressions (at an alpha of 0.05) and are not discussed further.

Next, we investigated whether the associations between time-use composition and the cognitive function outcomes were mediated by grey matter volume, frontal lobe volume, temporal lobe volume or hippocampus volume. For each cognitive outcome (long-term memory, executive function, processing speed), a series of linear regression models were fit, with each incorporating main effects of ROI volume and time-use composition, as well as the interaction between time-use composition and the respective ROI volume. All models were adjusted for covariates (age, sex, education).

Type II F-tests were used in determining variable significance (assessing variable effects after adjusting for other variables while adhering to the principle of marginality (44, 45)). To account for multiple comparisons, p-values within each of the final regression models were Benjamini-Hochberg false discovery rate adjusted (46, 47).

#### 2.4.3 Modelling reallocations of time

In the instance that 24-hour time-use composition was significantly associated with a volumetric outcome (following false discovery rate adjustment), we planned to plot model-generated predictive response curves to demonstrate how volumetric measures were associated with meaningful reallocations of time, using one for-remaining swaps (e.g., increasing MVPA by 20 minutes at the expense of all other behaviours equally) (25). Similarly, where an interaction between 24-hour time-use composition and a volumetric outcome was significantly associated with a cognitive outcome, we planned to plot model-generated predictive response curves to demonstrate how cognitive function was associated with meaningful reallocations of time across different brain volume levels (dichotomized to above or below the mean ROI volume) using one-for remaining swaps.

## 3. Results

### 3.1 Participant demographics

Of the original 426 participants recruited in the baseline phase of the ACTIVate study, 395 participants completed both T1 MPRAGE and T2 FLAIR imaging protocols. Seventeen participants were removed from the dataset as they did not have valid accelerometry data: 7 did not meet minimum criteria for a valid accelerometry dataset (i.e., less than minimum required days of recording); 8 were missing accelerometry data; and two had >1500 minutes of recorded time use per day. The overall final sample included 378 older adults (65.6 ± 3.0 years old, 123 males). Means, standard deviations and range (minimum and maximum) of key continuous variables are presented in Table 1. Participants had low white matter hyperintensity burden (mean = 2 ml) and were highly active, spending approximately 4.5 hours per day in physical activity (1.5 hours in MVPA; 3 hours in LPA), 11.1 hours in sedentary behaviour, and 8.4 hours sleeping. Participants’ time-use compositions are displayed in Figure 1.

**Table 1.**
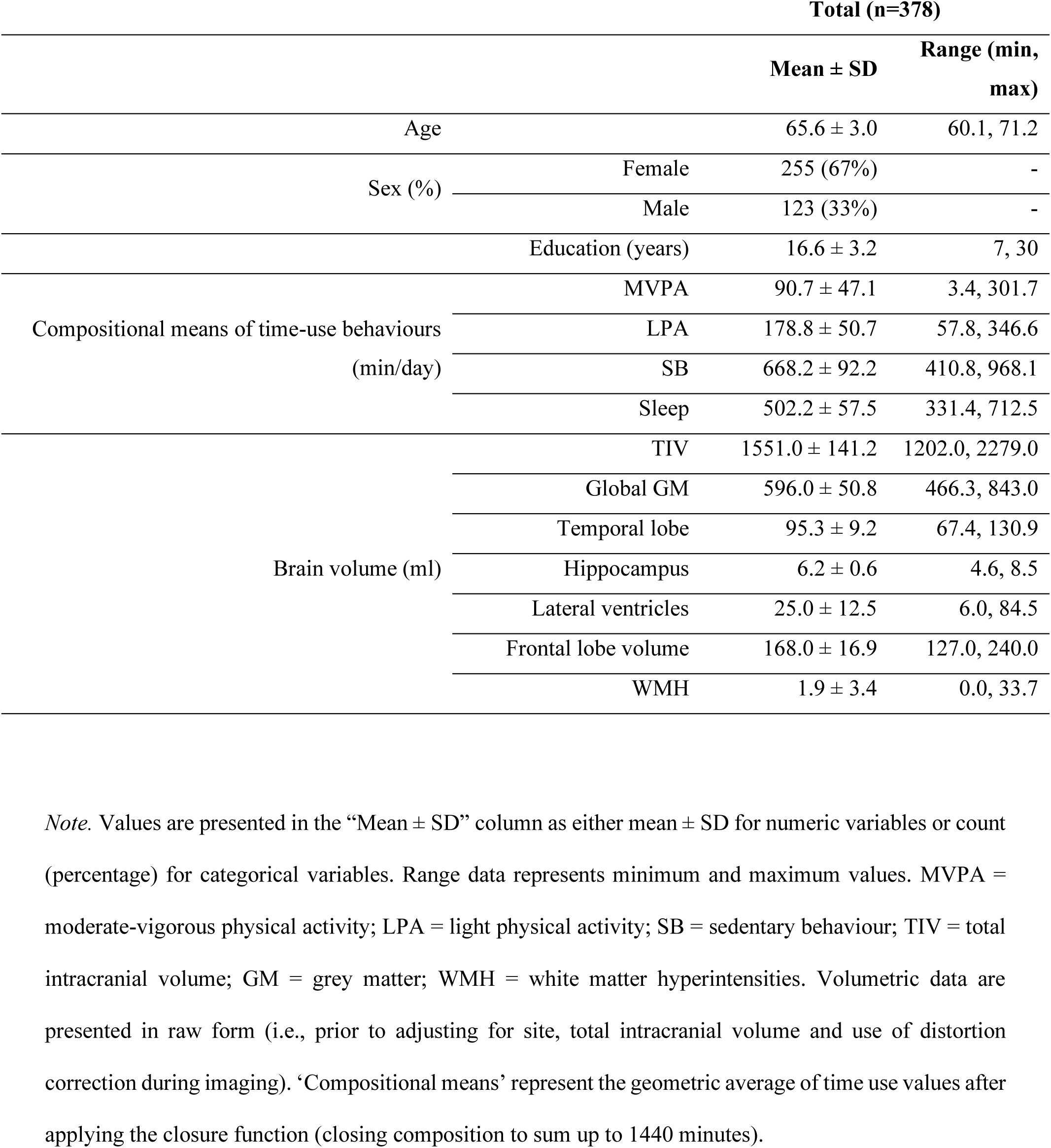
Participant demographics

**Figure 1.**
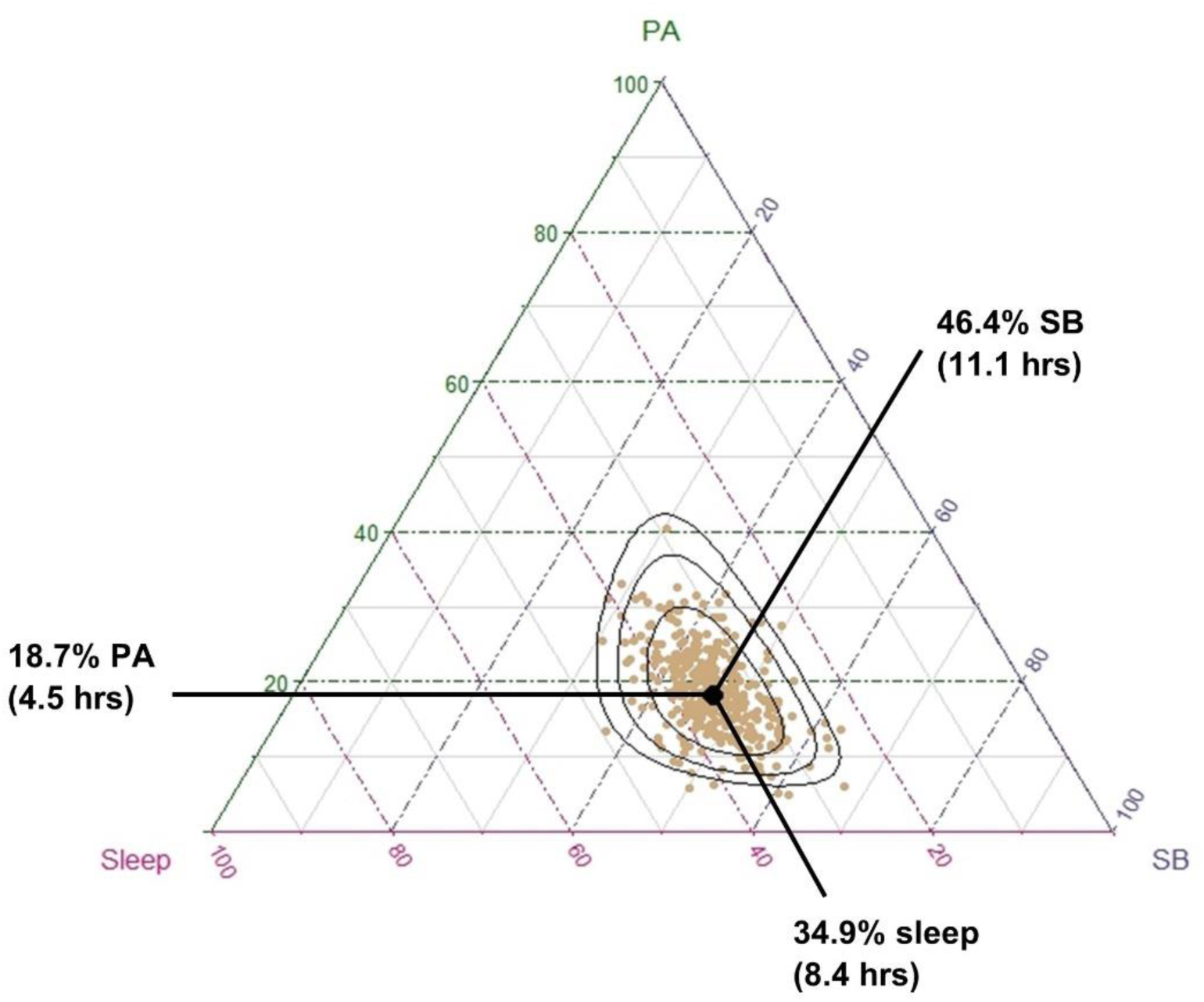
Distribution of participants’ time-use compositions. Each gold dot represents a single participant’s 24-hour time-use composition, whereas the black dot represents the average time-use composition of the entire sample (calculated as the compositional mean). On average, participants spent 18.7% of their day in physical activity (moderate-vigorous and light intensity, summed), 34.9% of their day in sleep, and 46.4% of their day in sedentary behaviour. Black ellipses represent 75%, 95% and 99% density contours assuming compositional normality (normal distribution on the simplex (42).

### 3.2 Associations between 24-hour time-use composition and brain structure

#### 3.2.1 Pairwise correlations

Pairwise Pearson correlations and their respective 95% confidence intervals are displayed in Table 2. There were no significant correlations between individual time-use behaviours and any volumetric outcomes. Several relationships were observed between individual time-use behaviours and cognitive outcomes: more time spent in MVPA was associated with faster processing speed (r= 0.18, 95% CI [0.07, 0.27]); more time spent in sedentary behaviour was associated with slower processing speed (r=-0.14, 95% CI [-0.24, -0.04]); and more time spent in sleep was associated with poorer long-term memory performance (r=-0.12, 95% CI [-0.23, -0.02]). Similarly, several ROI volumes were related to cognitive function: long-term memory was positively correlated with hippocampus volume (r=0.13, 95% CI [0.03, 0.23]); and executive function was positively correlated with total grey matter volume (r=0.22, 95% CI [0.12, 0.32]), temporal lobe volume (r=0.14, 95% CI [0.03, 0.24]), hippocampus volume (r=0.12, 95% CI [0.02, 0.22]) and frontal lobe volume (r=0.15, 95% CI [0.04, 0.25]) and negatively correlated with ventricle volume (r=-0.15, 95% CI [-0.25, - 0.05]). Higher age was associated with lower total grey matter (r=-0.28, 95% CI [-0.37, -0.19]), temporal lobe (r=-0.20, 95% CI [-0.29. -0.10]) and hippocampus volume (r=-0.16, 95% CI [-0.25, -0.06]), greater lateral ventricle volume (r=0.21, 95% CI [0.11, 0.30]), slower processing speed (r=-0.14, 95% CI [-0.24, - 0.04]) and worse executive function (r=-0.26, 95% CI [-0.35, -0.16]).

**Table 2:**
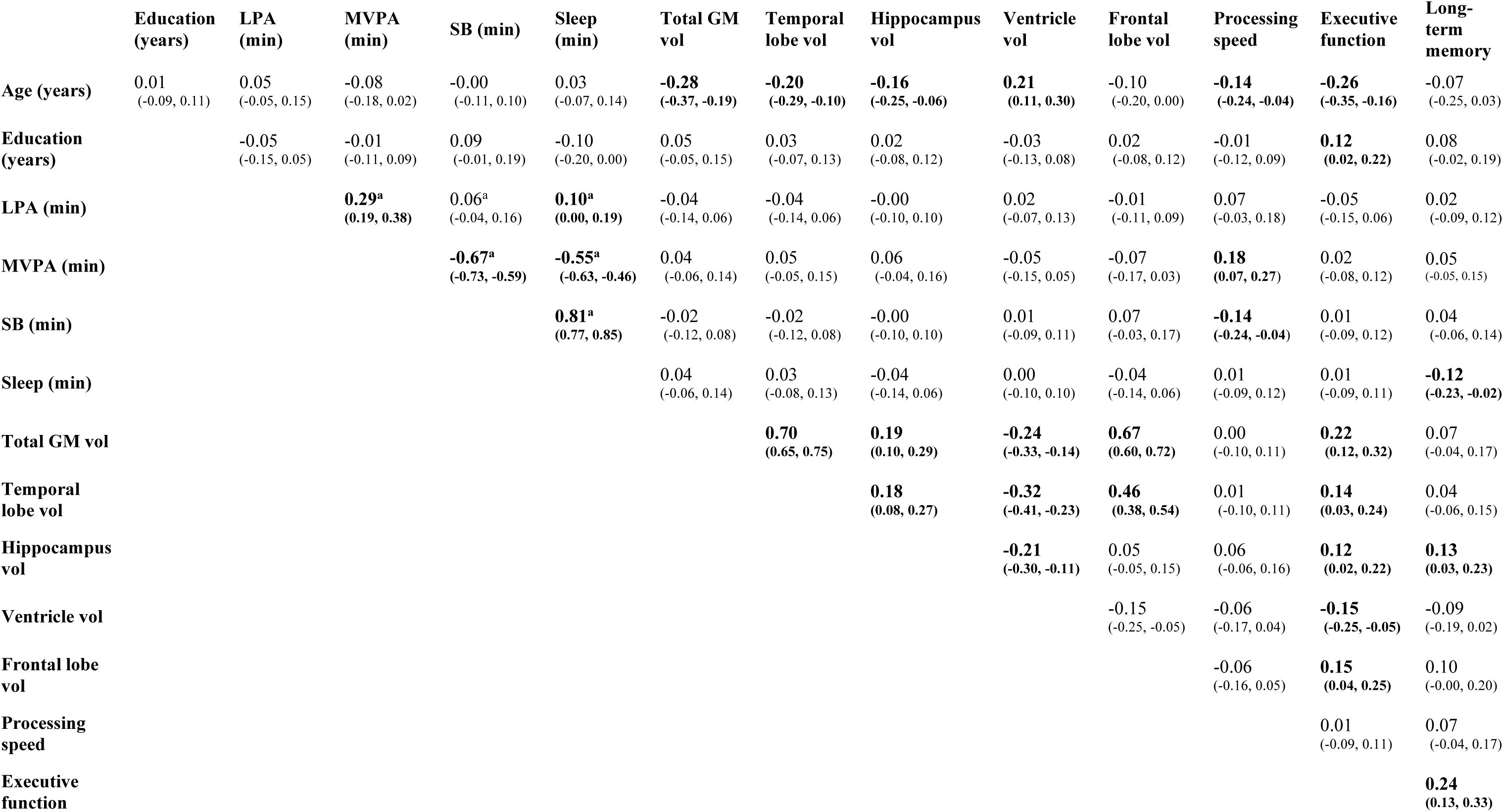
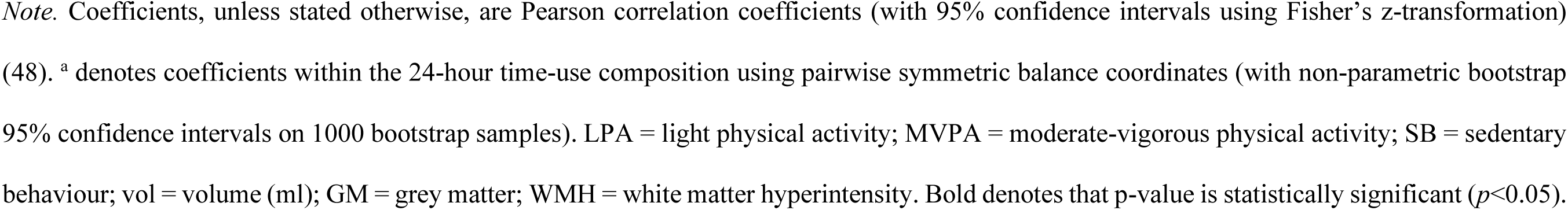
Pairwise correlations between continuous covariates, predictors, and outcome variables.

|  | Education<br>(years) | LPA<br>(min) | MVPA<br>(min) | SB (min) | Sleep<br>(min) | Total GM<br>vol | Temporal<br>lobe vol | Hippocampus<br>vol | Ventricle<br>vol | Frontal<br>lobe vol | Processing<br>speed | Executive<br>function | Long-term<br>memory |
| --- | --- | --- | --- | --- | --- | --- | --- | --- | --- | --- | --- | --- | --- |
| Age (years) | 0.01<br>(-0.09, 0.11) | 0.05<br>(-0.05, 0.15) | -0.08<br>(-0.18, 0.02) | -0.00<br>(-0.11, 0.10) | 0.03<br>(-0.07, 0.14) | **-0.28**<br>(-0.37, -0.19) | **-0.20**<br>(-0.29, -0.10) | **-0.16**<br>(-0.25, -0.06) | **0.21**<br>(0.11, 0.30) | -0.10<br>(-0.20, 0.00) | **-0.14**<br>(-0.24, -0.04) | **-0.26**<br>(-0.35, -0.16) | -0.07<br>(-0.25, 0.03) |
| Education<br>(years) |  | -0.05<br>(-0.15, 0.05) | -0.01<br>(-0.11, 0.09) | 0.09<br>(-0.01, 0.19) | -0.10<br>(-0.20, 0.00) | 0.05<br>(-0.05, 0.15) | 0.03<br>(-0.07, 0.13) | 0.02<br>(-0.08, 0.12) | -0.03<br>(-0.13, 0.08) | 0.02<br>(-0.08, 0.12) | -0.01<br>(-0.12, 0.09) | **0.12**<br>(0.02, 0.22) | 0.08<br>(-0.02, 0.19) |
| LPA (min) |  |  | **0.29<sup>a</sup>**<br>(0.19, 0.38) | 0.06<sup>a</sup><br>(-0.04, 0.16) | **0.10<sup>a</sup>**<br>(0.00, 0.19) | -0.04<br>(-0.14, 0.06) | -0.04<br>(-0.14, 0.06) | -0.00<br>(-0.10, 0.10) | 0.02<br>(-0.07, 0.13) | -0.01<br>(-0.11, 0.09) | 0.07<br>(-0.03, 0.18) | -0.05<br>(-0.15, 0.06) | 0.02<br>(-0.09, 0.12) |
| MVPA (min) |  |  |  | **-0.67<sup>a</sup>**<br>(-0.73, -0.59) | **-0.55<sup>a</sup>**<br>(-0.63, -0.46) | 0.04<br>(-0.06, 0.14) | 0.05<br>(-0.05, 0.15) | 0.06<br>(-0.04, 0.16) | -0.05<br>(-0.15, 0.05) | -0.07<br>(-0.17, 0.03) | **0.18**<br>(0.07, 0.27) | 0.02<br>(-0.08, 0.12) | 0.05<br>(-0.05, 0.15) |
| SB (min) |  |  |  |  | **0.81<sup>a</sup>**<br>(0.77, 0.85) | -0.02<br>(-0.12, 0.08) | -0.02<br>(-0.12, 0.08) | -0.00<br>(-0.10, 0.10) | 0.01<br>(-0.09, 0.11) | 0.07<br>(-0.03, 0.17) | **-0.14**<br>(-0.24, -0.04) | 0.01<br>(-0.09, 0.12) | 0.04<br>(-0.06, 0.14) |
| Sleep (min) |  |  |  |  |  | 0.04<br>(-0.06, 0.14) | 0.03<br>(-0.08, 0.13) | -0.04<br>(-0.14, 0.06) | 0.00<br>(-0.10, 0.10) | -0.04<br>(-0.14, 0.06) | 0.01<br>(-0.09, 0.12) | 0.01<br>(-0.09, 0.11) | **-0.12**<br>(-0.23, -0.02) |
| Total GM vol |  |  |  |  |  |  | **0.70**<br>(0.65, 0.75) | **0.19**<br>(0.10, 0.29) | **-0.24**<br>(-0.33, -0.14) | **0.67**<br>(0.60, 0.72) | 0.00<br>(-0.10, 0.11) | **0.22**<br>(0.12, 0.32) | 0.07<br>(-0.04, 0.17) |
| Temporal<br>lobe vol |  |  |  |  |  |  |  | **0.18**<br>(0.08, 0.27) | **-0.32**<br>(-0.41, -0.23) | **0.46**<br>(0.38, 0.54) | 0.01<br>(-0.10, 0.11) | **0.14**<br>(0.03, 0.24) | 0.04<br>(-0.06, 0.15) |
| Hippocampus<br>vol |  |  |  |  |  |  |  |  | **-0.21**<br>(-0.30, -0.11) | 0.05<br>(-0.05, 0.15) | 0.06<br>(-0.06, 0.16) | **0.12**<br>(0.02, 0.22) | **0.13**<br>(0.03, 0.23) |
| Ventricle vol |  |  |  |  |  |  |  |  |  | -0.15<br>(-0.25, -0.05) | -0.06<br>(-0.17, 0.04) | **-0.15**<br>(-0.25, -0.05) | -0.09<br>(-0.19, 0.02) |
| Frontal lobe<br>vol |  |  |  |  |  |  |  |  |  |  | -0.06<br>(-0.16, 0.05) | **0.15**<br>(0.04, 0.25) | 0.10<br>(-0.00, 0.20) |
| Processing<br>speed |  |  |  |  |  |  |  |  |  |  |  | 0.01<br>(-0.09, 0.11) | 0.07<br>(-0.04, 0.17) |
| Executive<br>function |  |  |  |  |  |  |  |  |  |  |  |  | **0.24**<br>(0.13, 0.33) |

*Note.* Coefficients, unless stated otherwise, are Pearson correlation coefficients (with 95% confidence intervals using Fisher's z-transformation) (48). <sup>a</sup> denotes coefficients within the 24-hour time-use composition using pairwise symmetric balance coordinates (with non-parametric bootstrap 95% confidence intervals on 1000 bootstrap samples). LPA = light physical activity; MVPA = moderate-vigorous physical activity; SB = sedentary behaviour; vol = volume (ml); GM = grey matter; WMH = white matter hyperintensity. Bold denotes that p-value is statistically significant ( $p < 0.05$ ).

#### 3.2.2 Associations between 24-hour time-use composition and brain volume

Table 3 displays outcomes of regression models testing the associations between 24-hour time-use composition and ROI volumes after adjusting for covariates (age, sex, education) and false discovery rate. In covariate-adjusted models, 24-hour time-use composition was not associated with total grey matter volume, frontal lobe volume, temporal lobe volume, hippocampal volume, or lateral ventricle volume both before and after adjusting for false discovery rate.

**Table 3.**
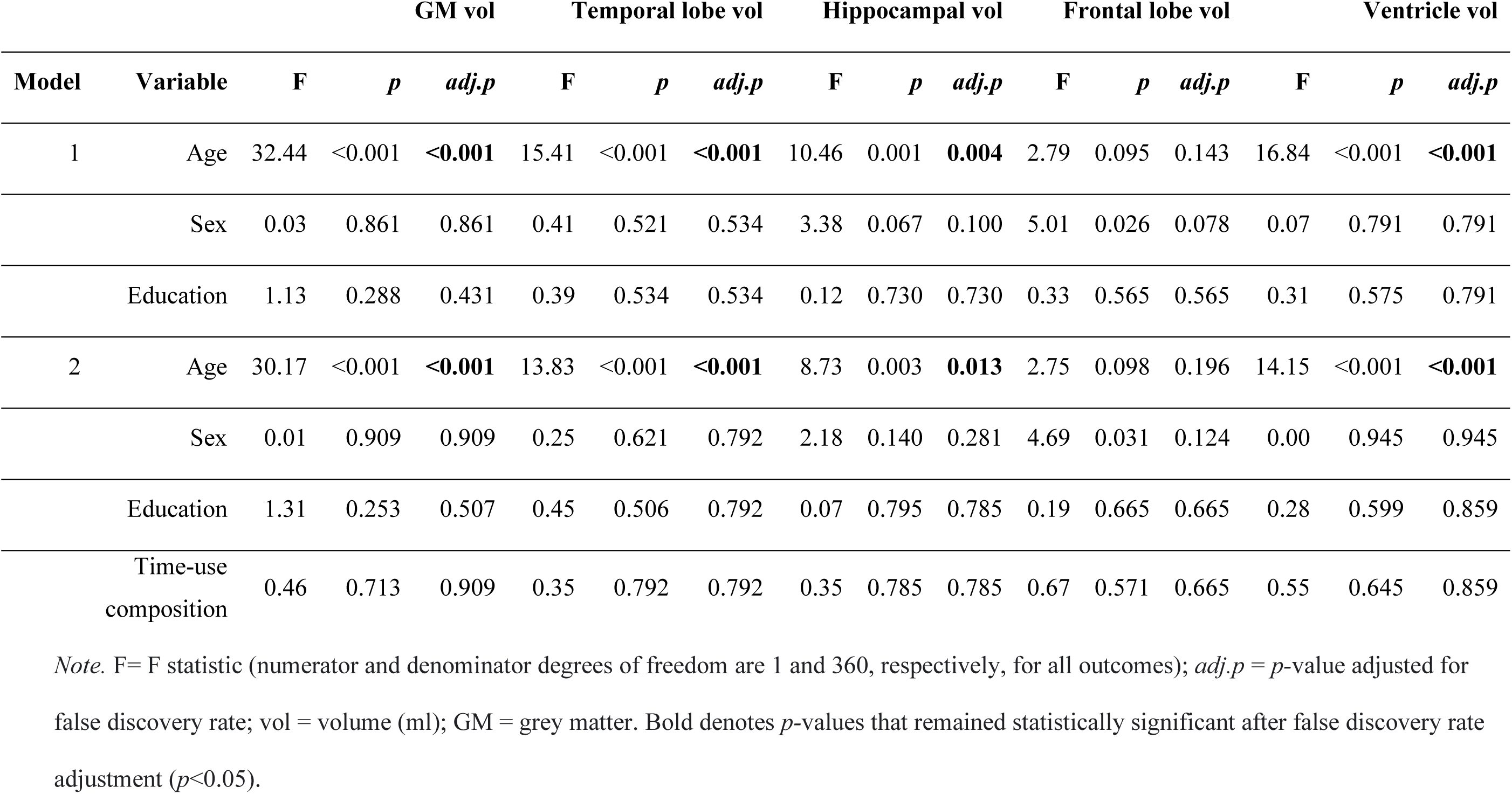
Statistical output of ANOVA Type II F-tests for volumetric outcomes.

| Model | Variable | GM vol |  |  | Temporal lobe vol |  |  | Hippocampal vol |  |  | Frontal lobe vol |  |  | Ventricle vol |  |  |
| --- | --- | --- | --- | --- | --- | --- | --- | --- | --- | --- | --- | --- | --- | --- | --- | --- |
|  |  | F | *p* | *adj.p* | F | *p* | *adj.p* | F | *p* | *adj.p* | F | *p* | *adj.p* | F | *p* | *adj.p* |
| 1 | Age | 32.44 | <0.001 | **<0.001** | 15.41 | <0.001 | **<0.001** | 10.46 | 0.001 | **0.004** | 2.79 | 0.095 | 0.143 | 16.84 | <0.001 | **<0.001** |
|  | Sex | 0.03 | 0.861 | 0.861 | 0.41 | 0.521 | 0.534 | 3.38 | 0.067 | 0.100 | 5.01 | 0.026 | 0.078 | 0.07 | 0.791 | 0.791 |
|  | Education | 1.13 | 0.288 | 0.431 | 0.39 | 0.534 | 0.534 | 0.12 | 0.730 | 0.730 | 0.33 | 0.565 | 0.565 | 0.31 | 0.575 | 0.791 |
| 2 | Age | 30.17 | <0.001 | **<0.001** | 13.83 | <0.001 | **<0.001** | 8.73 | 0.003 | **0.013** | 2.75 | 0.098 | 0.196 | 14.15 | <0.001 | **<0.001** |
|  | Sex | 0.01 | 0.909 | 0.909 | 0.25 | 0.621 | 0.792 | 2.18 | 0.140 | 0.281 | 4.69 | 0.031 | 0.124 | 0.00 | 0.945 | 0.945 |
|  | Education | 1.31 | 0.253 | 0.507 | 0.45 | 0.506 | 0.792 | 0.07 | 0.795 | 0.785 | 0.19 | 0.665 | 0.665 | 0.28 | 0.599 | 0.859 |
|  | Time-use composition | 0.46 | 0.713 | 0.909 | 0.35 | 0.792 | 0.792 | 0.35 | 0.785 | 0.785 | 0.67 | 0.571 | 0.665 | 0.55 | 0.645 | 0.859 |

*Note.* F= F statistic (numerator and denominator degrees of freedom are 1 and 360, respectively, for all outcomes); *adj.p* = *p*-value adjusted for false discovery rate; vol = volume (ml); GM = grey matter. Bold denotes *p*-values that remained statistically significant after false discovery rate adjustment (*p*<0.05).

#### 3.2.3 Cognitive function, brain volume and time-use composition

Regression outputs investigating interactions between 24-hour time-use composition and ROI volume for cognitive outcomes are displayed in Table 4. The interaction between time-use composition and total grey matter volume was associated with long-term memory and executive function outcomes prior to adjusting for false discovery rate. Similarly, the interaction between time-use composition and frontal lobe volume was associated with long-term memory and executive function outcomes. After false discovery rate adjustment, several interactions remained statistically significant: executive function was associated with the interaction between time-use composition and total grey matter volume (p_adj_=0.028) and the interaction between time-use composition and frontal lobe volume (p_adj_=0.018); and long-term memory remained significantly associated with the interaction between time-use composition and frontal lobe volume (p_adj_=0.018).

**Table 4.**
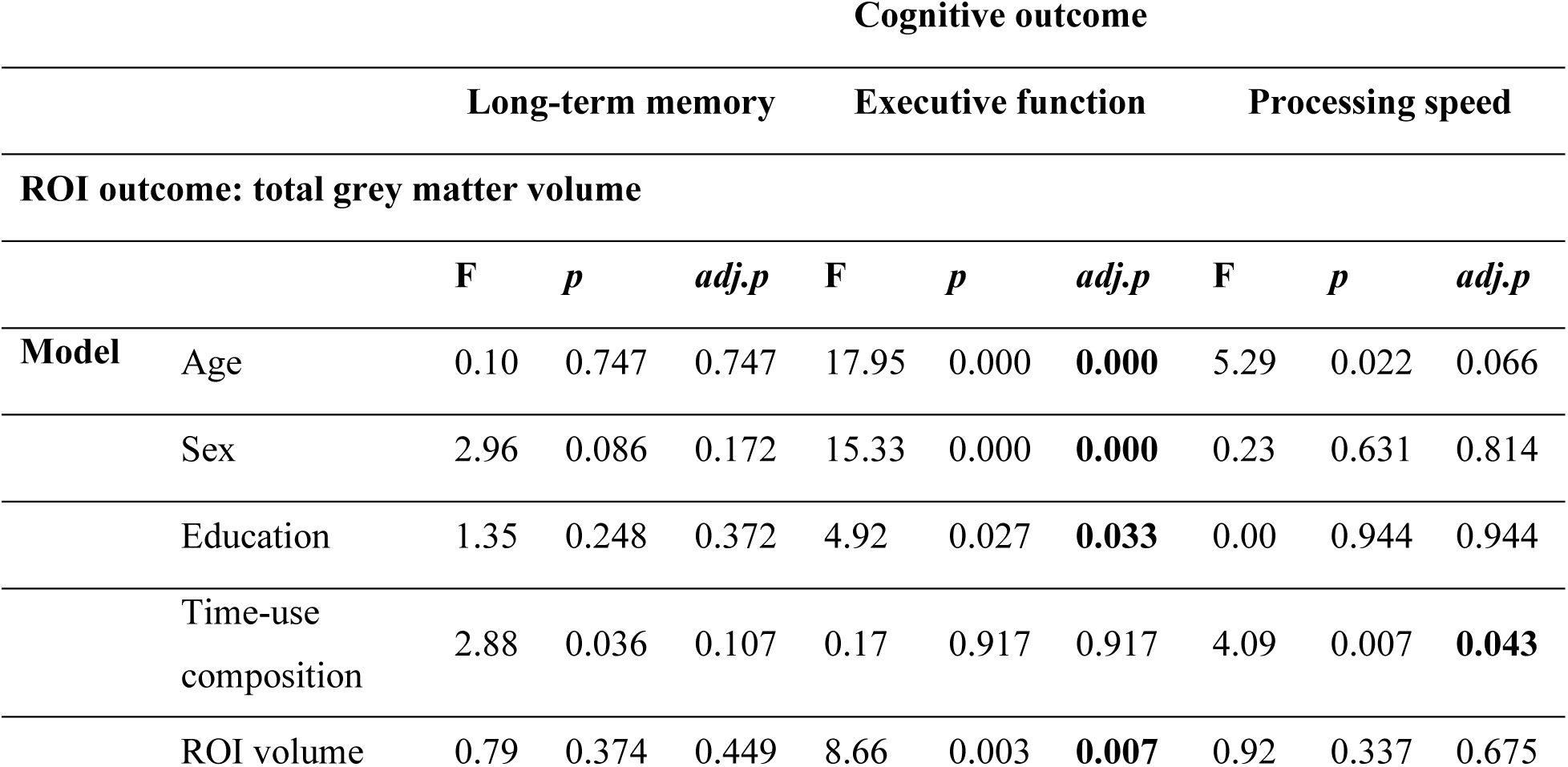
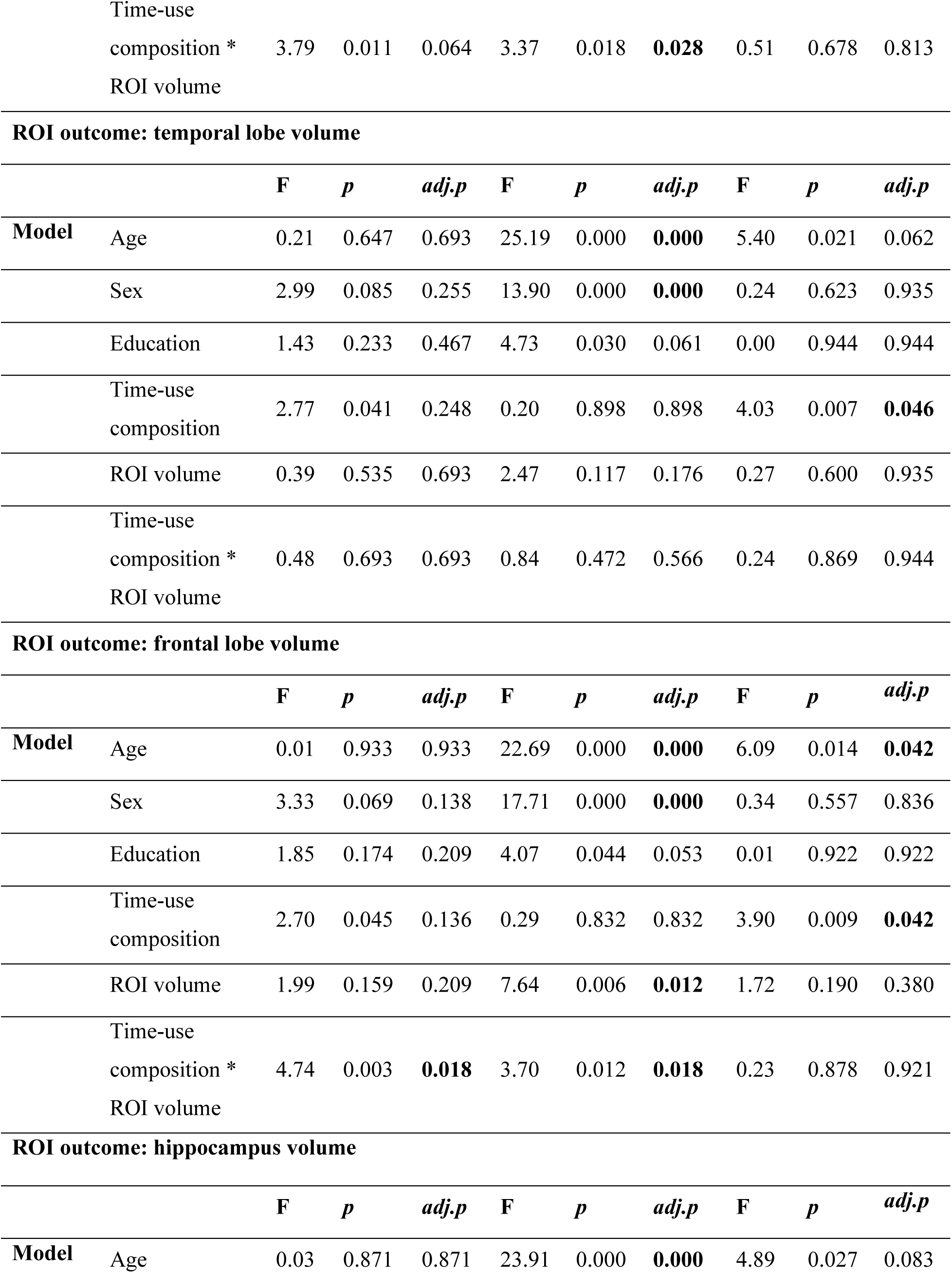
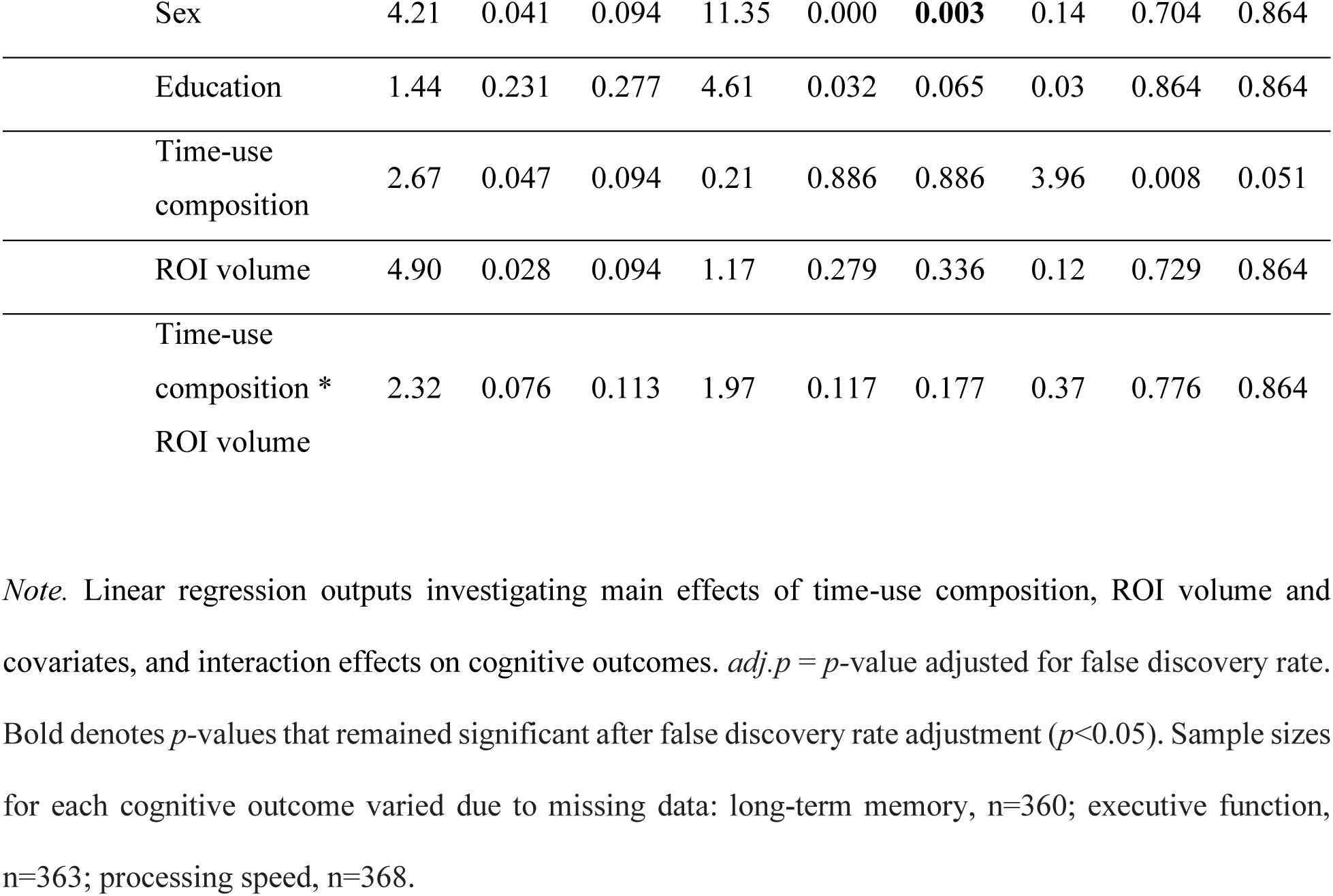
Linear regression outputs.

To further investigate these interactions, we plotted a series of model-estimated cognitive response curves which demonstrate the estimated associations of time reallocations with long-term memory and executive function outcomes, across high and low frontal lobe or total grey matter volumes, respectively. High and low volume groups were quantified as those above and below the mean frontal lobe volume (168ml) and total grey matter volume (596ml) in the sample, as the data were normally distributed (therefore a median split achieved similar data separation). Before creating the plots, regression models containing the significant interactions were replicated with frontal lobe volume and total grey matter volume included as categorical variables (two levels, upper and lower volume group, rather than as a continuous variable) to ensure that the interaction between time-use composition and each ROI volume remained significant. Interestingly, the interaction between time-use composition and frontal lobe volume (as a categorical variable) did not remain significant for executive function (p_adj_=0.80). This remained true when frontal lobe volume was split into quartiles (p_adj_=0.31). For this reason, the time-use composition by frontal lobe volume interaction for the executive function outcome was not further explored here. Figures 2 and 3 display the predicted differences in long-term memory z-score and executive function z-score associated with reallocations of time from the reference mean time-use composition towards and away from each time-use behaviour (positive and negative reallocations on the x-axis). Predicted associations are plotted separately for those above and below the mean frontal lobe and total grey matter volume, respectively.

**Figure 2:**
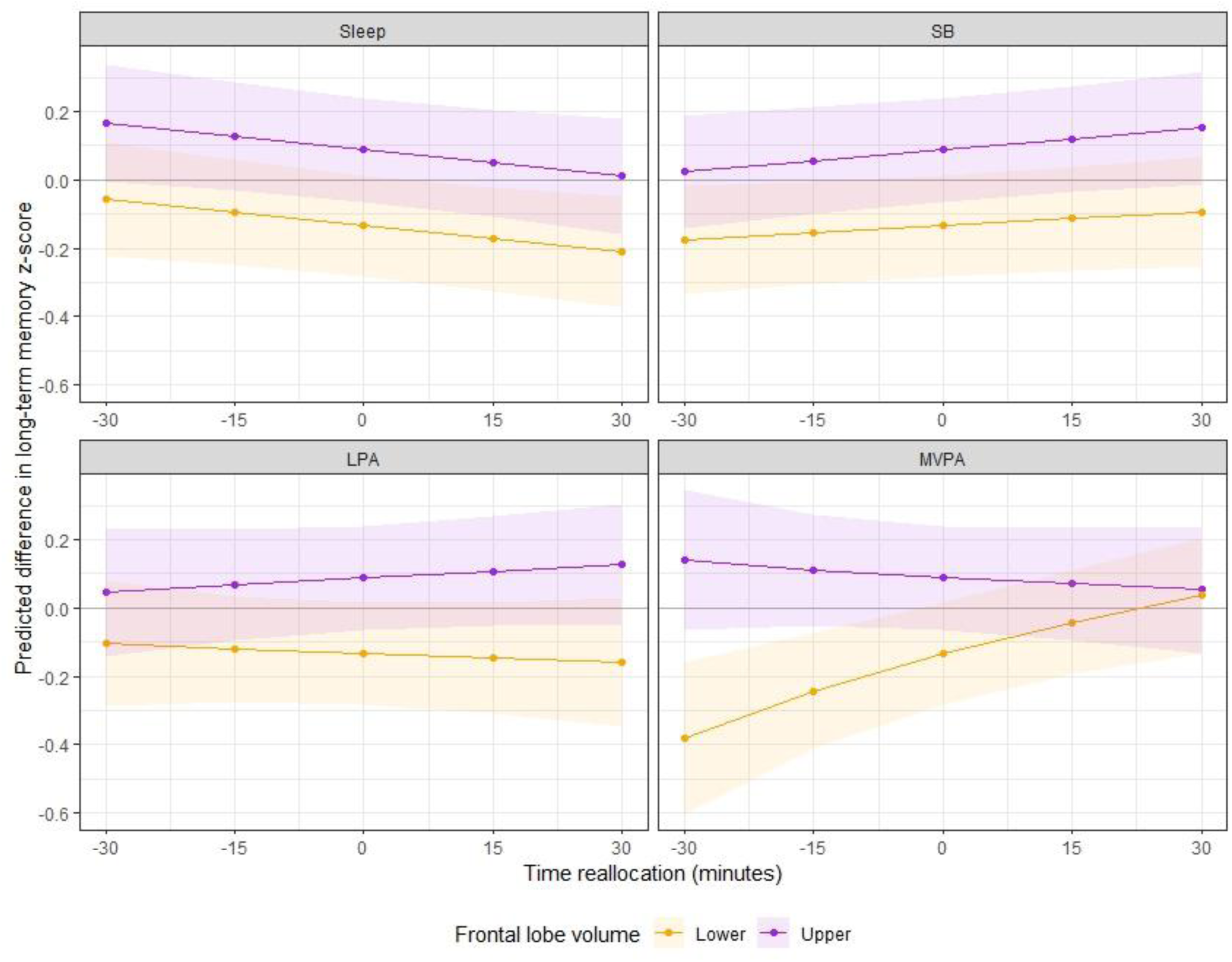
The model-predicted difference in long-term memory z-score (y-axis) associated with reallocations of time towards or away from each time-use behaviour (displayed in the header of each panel), in 15-minute increments from the reference mean time-use composition. Purple lines represent participants with greater than the sample mean frontal lobe volume (‘Upper’); orange lines represent participants with less than the sample mean frontal lobe volume (‘Lower’). Shading represents 95% confidence intervals. Mean frontal lobe volume (corrected) in the ‘upper’ group = 174.8 ± 5.0, range = 168.3, 193.0. Mean frontal lobe volume (corrected) in the ‘lower’ group = 161.2 ± 5.0, range = 143.3, 168.3.

**Figure 3:**
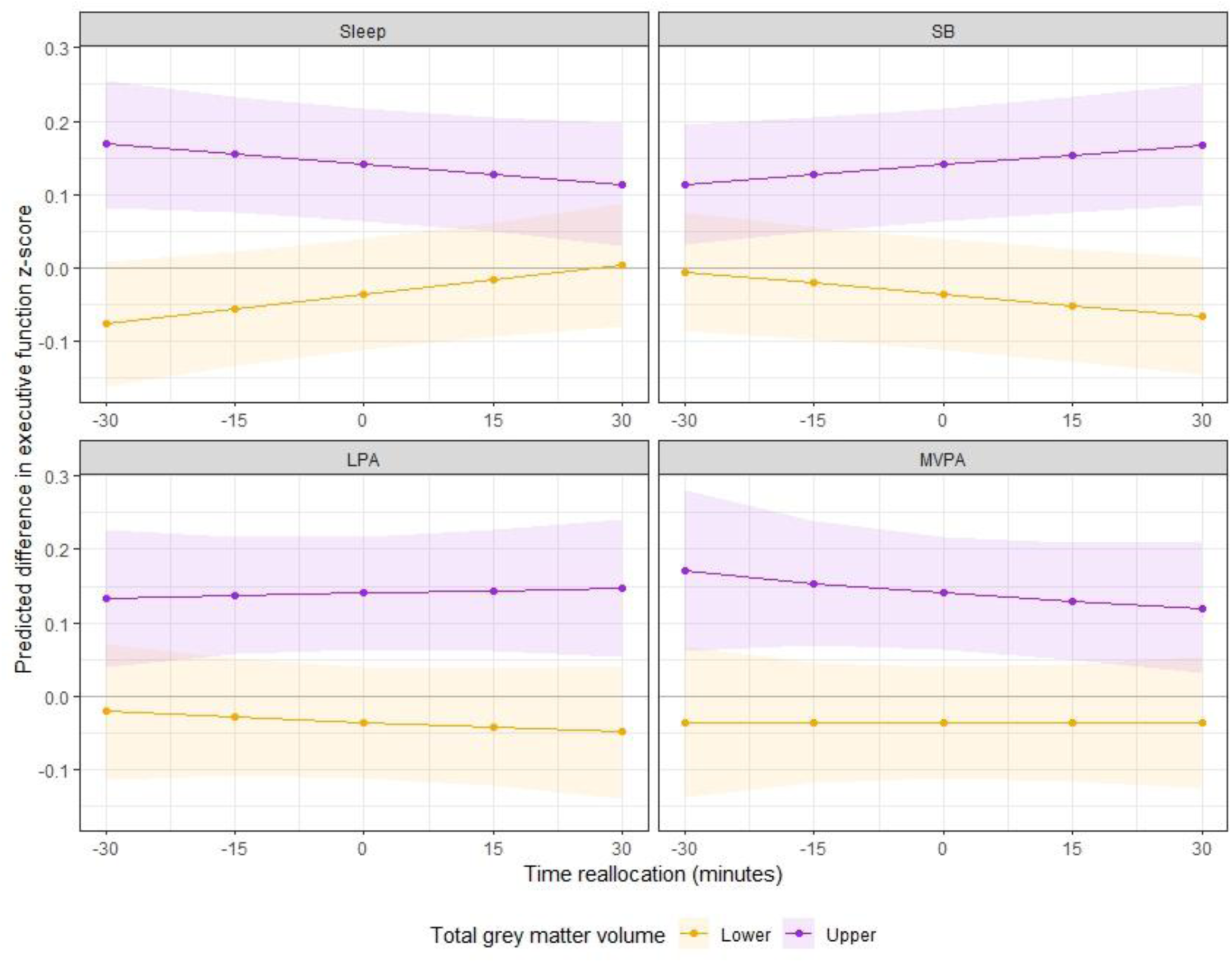
The model-predicted difference in executive function z-score (y-axis) associated with reallocations of time towards or away from each time-use behaviour (displayed in the header of each panel) in 15-minute increments from the reference mean time-use composition. Purple lines represent participants with greater than the sample mean total grey matter volume (‘Upper’); orange lines represent participants with less than the sample mean total grey matter volume (‘Lower’). Shading represents 95% confidence intervals. Mean total grey matter volume (corrected) in the ‘upper’ group = 611.4 ± 10.6, range = 596.7, 647.9. Mean total grey matter volume (corrected) in the ‘lower’ group = 580.7 ± 12.9, range = 519.8, 596.7.

Figure 2 suggests that for those with smaller frontal lobe volume (below the sample mean), more time in MVPA was favorably associated with long-term memory performance, whilst this time reallocation had little predicted benefit for those with greater frontal lobe volume (above the sample mean). More time in LPA was associated with small unfavorable differences in long-term memory performance for those with lower frontal lobe volume, and small favorable differences in performance for those with greater frontal lobe volume. Reallocating time towards or away from sleep and sedentary behaviour had similar (minimal) associations with long-term memory performance across both high and low frontal lobe volume groups.

Figure 3 shows that the executive function response curves for sleep and sedentary behaviour reallocations differ by total grey matter volume. For those with lower total grey matter volume, more time in sleep and less time in sedentary behaviour was favorably associated with executive function, while these reallocations were negatively associated with executive function in those with greater total grey matter volume. Reallocating time towards or away from LPA had minimal associations with executive function across both high and low total grey matter volume groups. Finally, spending more time in MVPA had small unfavorable associations with executive function in the higher total grey matter volume group, whilst reallocations towards or away from MVPA had minimal associations in the lower total grey matter group.

## 4. Discussion

### 4.1 Twenty-four-hour time-use composition and grey matter volume in healthy older adults

The primary aim of this study was to explore whether 24-hour time-use composition of MVPA, LPA, sedentary behaviour and sleep was associated with total and regional grey matter volume (temporal lobe, frontal lobe, hippocampus, and lateral ventricle volume) in healthy older adults. Our main finding was that there were no associations between 24-hour time-use composition and any volumetric outcomes. This finding is likely not a result of accounting for all four time-use behaviours together (i.e., the effects of one time-use behaviour “canceling out” or altering the effect of another) as there were no significant pairwise correlations between independent time-use behaviours with any volumetric outcomes. As this study was the first to investigate associations between 24-hour time-use composition and grey matter volume, it is challenging to contextualize the findings with previous research. However, the findings of our correlation analyses contradict several previous studies in older adults which did report significant associations between grey matter volume and physical activity (4, 6, 7, 11), sedentary behaviour (15) or sleep (19, 20) when considered independently.

The discrepancies between our study and previous reports may be due to several factors. First, the sample used in the current study was recruited for a longitudinal study which aims to map differences in lifestyle (time use and diet) against changes in cognitive function, therefore at baseline participants were required to be healthy with no cognitive impairment. Despite best efforts to recruit participants across diverse lifestyle profiles, the sample had high cognitive function (mean score on Addenbrooke’s Cognitive Examination III at baseline was approximately 95/100 in the ACTIVate sample (27)), participants were highly active (mean total PA per day = 4.5hrs), and there was little variability in time-use composition at baseline across the sample (see Figure 1). Conversely, several studies which did observe associations between physical activity or sleep and grey matter volume were conducted in populations with greater variability in time use profiles (4, 9). For example, Bugg and Head (9) reported that physical exercise engagement (measured as self-reported engagement in running, walking or jogging over the past 10 years) was associated with volume in the superior frontal cortex. Due to extreme skewness in their data, participants were categorized in to low or high engagement groups, whereby the mean levels of physical exercise engagement were 0.63 and 7.76, respectively (sample range = 0-29.72). The authors reported that those in the low engagement group engaged in moderate intensity activity less than 2 days per week for 10 minutes, whereas the average amount of moderate activity for the high engagement group was approximately 5 days per week for 30 minutes each (9). Similarly in a longitudinal study, Tan and colleagues (4) reported that Physical Activity Index (PAI) scores at baseline were significantly associated with total cerebral and hippocampal volumes at follow-up (∼10 years). PAI scores were generated by asking participants to report the number of hours per day spent sleeping (weighting factor = 1), sedentary (weighting factor = 1.1), or in light (weighting factor = 1.5), moderate (weighting factor = 2.4) and heavy activities (weighting factor = 5). Thus, a score of 24 reflects a day spent sleeping continually (4). The range of PAI scores in their sample ranged from 24.2-65.7 in females, and 24.6-74.5 in males, with median PAI scores of 34.4 and 35.4, respectively. On balance, a much wider range of time-use profiles were included in their study and it could be that the observed relationships between volumetric outcomes and activity levels were driven by extreme cases (e.g., highest quintiles relative to lowest) (4). Similarly, other studies that have reported associations between physical activity or sleep and grey matter volume observed differences between extreme sub-groups only (i.e., lowest VS. highest quintile of physical activity) (6, 19).

Although there are few studies investigating associations between sedentary behaviour and grey matter volume in older adults, the null findings of the our study align with that of a recent review by Maasakkers et al. (18) who reported inconclusive evidence for the relationship between sedentary behaviour and grey matter volume. As hypothesized by Maasakkers et al. (18), it may be that white matter volume is more sensitive to the physiological effects of sedentary behaviour compared to grey matter. It should also be noted that several other studies have reported no significant associations between sedentary behaviour and total grey matter volume (in line with the current study), but did find significant differences between low and high sedentary behaviour groups in smaller sub-regions, such as hippocampal sub-fields (14, 15). Given that we found no association between sedentary behaviour and total hippocampal volume in this study, it may be possible that the ROIs investigated in this study were not sensitive enough to detect differences which may be present in sub-regions.

Another key difference between the current study and previous studies which may contribute to discrepancies in findings is the age of the sample. Participants in the ACTIVate study were aged 60-70 years at baseline (mean age = 66 ± 3 years), a narrow age range chosen in order to capture healthy older adults who may be in pre-clinical stages of dementia (i.e., where brain changes are beginning to occur with no behavioural or cognitive symptoms). Several studies that have reported associations between independent time-use behaviours and grey matter volume were conducted in older samples (e.g., 70-80 years) which may have more progressed brain atrophy in comparison (4, 7), or in samples with a wider age range (e.g., including middle-age and older adults in sample). It is plausible that the observed differences in these studies may have been driven by older participants.

Finally, as is typical for time-use research, there are considerable inconsistencies in the measures used to estimate physical activity, sedentary behaviour, and sleep across studies (i.e., subjective recall versus accelerometry), which limits comparability with the current study. To our knowledge, the current study was the first to assess 24-hour time-use composition against grey matter volume in older adults using accelerometry, whereby participants’ activity patterns were monitored by a wrist-worn device for 24-hours per day. Comparatively, previous studies which have reported relationships between independent time-use behaviours and grey matter volume outcomes have mostly used self-report measures of duration (19) or validated questionnaires such as the Pittsburgh Sleep Quality Index (20), PAI (4), or activity compendiums (11). Moreover, studies which have used accelerometry to derive activity patterns have not taken a 24-hour approach, and have quantified physical activity outcomes using different metrics including total physical activity (irrespective of intensity) (7) or classification into low and high levels of MVPA (6). Taken together, the variability in measures used to capture activity patterns, as well as the absence of studies that have taken a 24-hour compositional approach, limits the comparability of the current study to previous studies.

### 4.2 Grey matter volume as a mechanism linking 24-hour time use and cognitive function

Several mechanisms likely underlie the relationship between lifestyle and cognitive function in older adults, including maintenance of grey matter volume. This relationship has only been investigated when considering physical activity, sedentary behaviour and sleep independently, rather than as a 24-hour composition. In our secondary analysis, we found that long-term memory was associated with the interaction between 24-hour time-use composition and frontal lobe volume, and that executive function was associated with the interaction between 24-hour time-use composition and both total grey matter volume and frontal lobe volume (although, the frontal lobe by executive function interaction was not further explored).

Predictive modelling allowed further understanding of these interactions while separating participants by ROI volume (above or below the sample mean). For long-term memory outcomes, reallocating time towards or away from LPA, sedentary behaviour or sleep (at the equal expense of all other behaviours) had minimal predicted associations with performance regardless of frontal lobe volume. However, reallocating time towards or away from MVPA was associated with long-term memory performance, and this relationship was more pronounced for those with smaller frontal lobe volume. The same reallocation, e.g., spending 30 minutes less time in MVPA, was associated with a ∼0.25 SD lower long-term memory z-score for those with smaller frontal lobe volume, but only a slight difference (∼0.05 SD higher) in long-term memory z-score for those with larger frontal lobe volumes. This finding suggests that benefits from spending more time in MVPA (or more so, the deficits from reducing time spent in MVPA) on memory performance may depend on frontal lobe volume. There are several key points that could be considered to contextualize these findings. First, evidence suggests that performing physical activity at higher intensities is positively associated with concentrations of neurochemicals which may enhance memory and learning (49, 50). Second, evidence from both animal and human studies suggests that effects of physical activity on the brain appear to be specific, in that particular brain regions are more sensitive to the benefits of physical activity than others (51). One such brain region is the frontal cortex, which along with the hippocampus, also typically shows marked age-related atrophy compared to other regions (51). On balance, it has been suggested that brain regions with more age-related atrophy may yield the most benefits from physical activity (51). This may explain why the predicted impact of increasing or decreasing time in MVPA on long-term memory performance was more prominent in participants with smaller frontal lobe volumes.

Interestingly, our secondary analysis did not infer the same importance for MVPA in those with lower total grey matter volume in the context of executive function outcomes. Reallocating time towards or away from LPA or MVPA had minimal predicted associations with performance regardless of total grey matter volume (increasing MVPA had slight unfavorable associations in those with greater volume). However, reallocating time towards sleep or away from sedentary behaviour appeared favorably associated with executive function (+0.03SD for each 30-minute reallocation) in older adults with smaller total grey matter volume, whilst the opposite was true for those with greater total grey matter volumes (less sleep and more sedentary behaviour = better executive function). Whilst these findings somewhat align with a recent study by Tai et al. (52) who suggested brain volume mediates the relationship between sleep duration and executive function, they are difficult to contextualize due to the cross-sectional nature of the study and the novelty of our analytical approach. We propose several potential contributors to these findings. For example, it is possible that those with smaller grey matter volume were achieving good *quality* sleep at baseline, so increasing time in this behaviour would be beneficial for executive function (whilst the opposite may have been true for those with greater volume). Alternatively, it may be that those with smaller total grey matter volume were engaging in sedentary behaviours which are not beneficial for cognitive function (i.e., TV watching), whilst those with greater total grey matter volume were engaging in cognitively stimulating sedentary behaviours (i.e., computer use, reading), so reallocating time away from or towards sedentary behaviour, respectively, would be beneficial for executive function (16). It should be noted that a previous study found no impact of sedentary behaviour context or sleep quality ratings on associations between 24 hour time-use composition and executive function in the ACTIVate cohort, but these analyses were conducted in the total sample only (and were not stratified by grey matter volume) (27). Above all, it is important to note that the predicted differences in executive function resulting from each reallocation in this study were much smaller compared to those observed for long-term memory, so these findings should be interpreted with caution. This is likely because the range in executive function z-scores in this cohort were much narrower (range = -1.77, 1.14) than long-term memory z-scores (range = -3.59, 1.58), so smaller differences in executive function z-score (resulting from each reallocation) were likely needed to achieve statistical significance.

Importantly, results in these secondary analyses were derived from cross-sectional data and the predicted associations were modest in scale, and therefore should be interpreted with some caution. Despite correcting each ROI volume for total intracranial volume in this study, we cannot deduce that smaller ROI volumes reflect accelerated atrophy due to the cross-sectional nature of the study. If each of these findings are upheld in longitudinal studies, they may indicate that lifestyle interventions which aim to maintain or improve cognitive functions such as memory and executive function (as a means to reduce dementia risk or delay dementia onset) could be tailored based on individual differences in brain volume (i.e., prescribing higher intensity physical activity for those with more progressed brain atrophy in the frontal lobe, or targeting sleep duration in those with more progressed total grey matter atrophy).

### 4.3 Strengths, limitations, and future directions for research

The current study is the first to investigate associations between 24-hour time use and grey matter volume using a compositional data analysis approach. We used reliable cognitive tests and device-based measures of time use which may be less susceptible to recall bias in older adults, and took a conservative approach to our data analysis by accounting for technical differences in scanning across the cohort (scanner type and use of distortion correction function) and by adjusting for false discovery rate which is not typically done in exploratory research (47). There are several limitations that should be noted. As outlined in a previous study using the same dataset (27), the recruited sample were highly active and highly educated despite best efforts to recruit participants across a variety of activity and dietary patterns. The cross-sectional nature of the study limits the inferences that can be made about causal relationships between variables.

## 5. Conclusions

The current study found no associations between 24-hour time-use composition and measures of global and regional grey matter volume in a sample of healthy older adults without dementia. We found some evidence that grey matter volume (globally, and in the frontal lobe) may mediate the relationship between 24-hour time-use composition and specific cognitive functions. These relationships should be explored longitudinally and in a more diverse sample to better understand the directionality and temporal effects of 24-hour time use on grey matter volume and cognitive function in healthy older adults.

## Data Availability

All data produced in the present study are available upon reasonable request to the authors.

## List of abbreviations

*MRI =*: magnetic resonance imaging
*METs =*: metabolic equivalents
*MVPA =*: moderate-vigorous physical activity
*LPA =*: light physical activity
*SB =*: sedentary behaviour
*MPRAGE =*: magnetization-prepared rapid gradient-echo
*FLAIR =*: fluid-attenuated inversion recovery
*CANTAB =*: Cambridge Neuropsychological Test Automated Battery
*CoDA =*: compositional data analysis
*ROI =*: region of interest
*PAI =*: Physical Activity Index
*SD =*: standard deviation

## Declarations

### Ethics approval and consent to participate

Ethics approval was obtained from the University of South Australia and University of Newcastle Human Research Ethics Committee (202639).

### Consent for publication

Not applicable.

### Availability of data and materials

The datasets generated and/or analysed during the current study are not publicly available due to the longitudinal nature of the larger ACTIVate study (ongoing at the time of publication) but are available from the corresponding author on reasonable request.

### Authors’ contributions

MM contributed to the study design, data analysis, interpretation of results, writing, and drafting of the manuscript. DD, TS, JF and YX contributed to the data analysis, interpretation of results, and drafting of the manuscript. AW and FK contributed to the study design, interpretation of results, and drafting of the manuscript. TO, MG, JD and MB contributed to interpretation of results and drafting of the manuscript. AS contributed to study design, data collection, interpretation of results and drafting of the manuscript. All authors contributed to the article and approved the submitted version.

## Acknowledgements

We firstly thank the imaging teams at the Hunter Medical Research Institute and the Clinical Research Imaging Centre (SAHMRI site) for your invaluable assistance in collecting MRI data for our study. We thank Montana Hunter, Louise Massie, Kate Dyer, Johanna Paddick, Felicity Simpson, Ashlee Harvey, Nicholas Ware, Leon Day, Elena Milochis, Alannah Graziano, Elizabeth Taddeo, Karen Wilson, Jenna Johnson, Nathan Tran, Gemma Mieko Furuhashi, Helen Nicholas, Riley Jackson, Teigan Cotterill, Mahmoud Abdolhoseini, Fayeem Aziz, and David Metherell for their valued contribution towards data collection and study coordination. We also thank the HMRI Research Volunteer Register for partial recruitment of participants at the Newcastle site.

